# Population based estimates of comorbidities affecting risk for complications from COVID-19 in the US

**DOI:** 10.1101/2020.03.30.20043919

**Authors:** Mary L. Adams, David L. Katz, Joseph Grandpre

## Abstract

We used 2017 Behavioral Risk Factor Surveillance System (BRFSS) data (N=444,649) to estimate the proportion of US adults who report comorbidities that suggest heightened risk of complications from COVID-19. Co-morbidities included cardiovascular disease, chronic obstructive pulmonary disease (COPD), diabetes, asthma, hypertension, and/or cancer other than skin, based on data from China. Overall 45.4% (95% CI 45.1-45.7) of adults reported any of the 6 comorbidities, increasing from 19.8% (19.1-20.4) for ages 18-29 years to 80.7% (79.5-81.8) for ages 80+ years. State rates ranged from 37.3% (36.2-38.5) in Utah to 58.7% (57.0-60.4) in West Virginia. Rates also varied by race/ethnicity, health insurance status, and employment. Excluded were residents of nursing homes or assisted living facilities. Although almost certainly an underestimate of all adults at risk due to these exclusions, these results should help in estimating healthcare needs for adults with COVID-19 complications living in the community.

**Article Summary Line:** Overall, 45.4% of US adults were estimated to be at heightened risk of COVID-19 complications due to co-morbidities, increasing from 19.8% for ages 18-29 years to 80.7% for ages 80+ years, with state-to-state variation.

## Introduction

Since COVID-19 was discovered in China in late 2019, cases have spread far and wide and are putting a strain on healthcare systems in many countries. Data from China indicate that while 81% of patients had mild cases, 14% of cases were severe and 5% were critical. (*1,2*). The overall case fatality rate (CFR) in China for 55,924 confirmed cases was 3.8% (*3*) with increased rates in adults with comorbid conditions including cardiovascular disease (CVD; CFR=13.2%, diabetes (9.2%), chronic respiratory disease (8.0%), hypertension (8.4%), and cancer (7.6%); the case fatality rate for those with none of these comorbid conditions was 1.4% (*3*). Our objective for this study was to use population based US data to estimate the approximate fraction of adults in the community who might be at heightened risk for complications from COVID-19 because they reported one of the 6 chronic conditions with high CFR in China.

## Methods

We used publicly available 2017 Behavioral Risk Factor Surveillance System (BRFSS) data (*4*) from telephone surveys of 444,649 randomly selected adults ages 18 and older in the 50 states and the District of Columbia (DC). The BRFSS includes only non-institutionalized adults so residents of nursing homes and assisted living facilities are among those not surveyed. We chose to use 2017 data in order to include hypertension which was not addressed in 2018. Data were adjusted for the probability of selection and weighted to be representative of the adult population in each state by age, gender, race/ethnicity, marital status, education, home ownership, and telephone type and included weights and stratum variables needed for analysis. The median response rate for cell phone and land line surveys combined was 47.2%, ranging from 33.9% in California to 61.1% in Utah (*5*). In addition to residents of nursing homes or assisted living facilities, persons unable to respond to a telephone survey are also excluded during the random selection process within a household. Reliability and validity of the BRFSS have been found to be moderate to high for many survey measures, in particular those used here which can be checked versus medical records (*6*).

### Measures

The key variable was a composite measure including adults reporting they were ever told they had cardiovascular disease (CVD: a heart attack, angina, coronary heart disease, or a stroke), diabetes, current asthma (included as a chronic respiratory disease (*2*)), chronic obstructive pulmonary disease (COPD), hypertension, and/ or cancer other than skin. The number of comorbid conditions was counted for each respondent and the proportion of all adults who reported at least one of the comorbid conditions was considered to represent adults at heightened risk of complications from COVID-19. A secondary measure was receipt of a seasonal influenza vaccination in the past year as a rough estimate of potential demand for a COVID-19 vaccine when available.

Demographic measures included age group (18-29, 30-39, 40-49, 50-59, 60-69, 70-79, and 80+ years, which was created by combining 5 year age groups provided in the data set), self-reported race/ethnicity (non-Hispanic white, Black or African American, Hispanic of any race, American Indian/Alaska native, Asian/Pacific Islander, and other), health insurance coverage (any kind of health care coverage, including health insurance, prepaid plans such as HMOs, or government plans such as Medicare, or Indian Health Service), employment status (employed or self-employed, out of work, homemaker, student, retired, or unable to work), and state of residence which included the District of Columbia.

### Analysis

Stata version 14.1 (Stata Corp LP, College Station, TX) was used for data analysis to account for the complex sample design of the BRFSS. Point estimates and 95% confidence intervals are reported using the landline and cell phone weights, plus stratum and PSU variables supplied in the data set (*4*). Missing values were excluded from analysis. A total of 11,508 records from the original survey sample of 444,649 were excluded due to missing data for any of the 6 chronic conditions, leaving a total N of 433,141. An additional 5,694 records were excluded due to missing values for age resulting in a final N for data including age groups of 427,447.

We also used available case fatality rates from China (*3*) to estimate the fraction of adults in that country with any of the 6 comorbid conditions. We used the formula below and different estimates of the CFR for adults with at least one of the comorbid conditions, ranging from 7.6% - 13.2%, including 9.28% which was the average, and 8.4% which is the rate for hypertension, the most prevalent comorbid condition (*3*); 1.4% was the CFR for those with none of the 6 and 3.8% was the overall CFR.

Formula: (CFR *X) + (1.4*(1-X)) =3.8 where CFR = rate of risk group and X= % in that risk group.

## Results

Among all 444,649 survey respondents, 48.7% were male, 13.9% were ages 70+, 63.3% were white, 18.2% were retired, and 12.1% were uninsured, with similar results for the study sample with 11,508 records with missing values removed. Overall 45.4% (95% CI 45.1-45.7) of respondents reported one or more of the 6 comorbidities and were considered at heightened risk for complications from COVID-19. Among all adults, 26.7% (26.5-27.0) reported 1 co-morbidity, 12.0% (11.8-12.2) reported 2, 4.7% (4.6-4.8%) reported 3 and 2.0% (1,9-2.1) reported 4 or more. Prevalence rates of the separate comorbidities were 8.5% for CVD, 6.6% for COPD, 9.1% for asthma, 10.8% for diabetes, 32.4% for hypertension, and 6.8% for cancer. Rates of reporting one or more of the comorbid conditions increased from 19.8% (19.1-20.4) for ages 18-29 years to 80.7% (79.5-81.8) for ages 80+ years (Table 1.) While the percentage of adults with any of the comorbidities increased with age, over half (53.4%) of the total were ages 18-59 years because there are more adults in that age range compared with those 60 and older (71.3% vs. 28.7% respectively). State rates ranged from 37.3% (36.2-38.5) in Utah to 58.7% (57.0-60.4) in West Virginia. Rates also varied by self-reported race/ethnicity, health insurance status, and employment, but not by gender (Table 1).

**Table 1.**
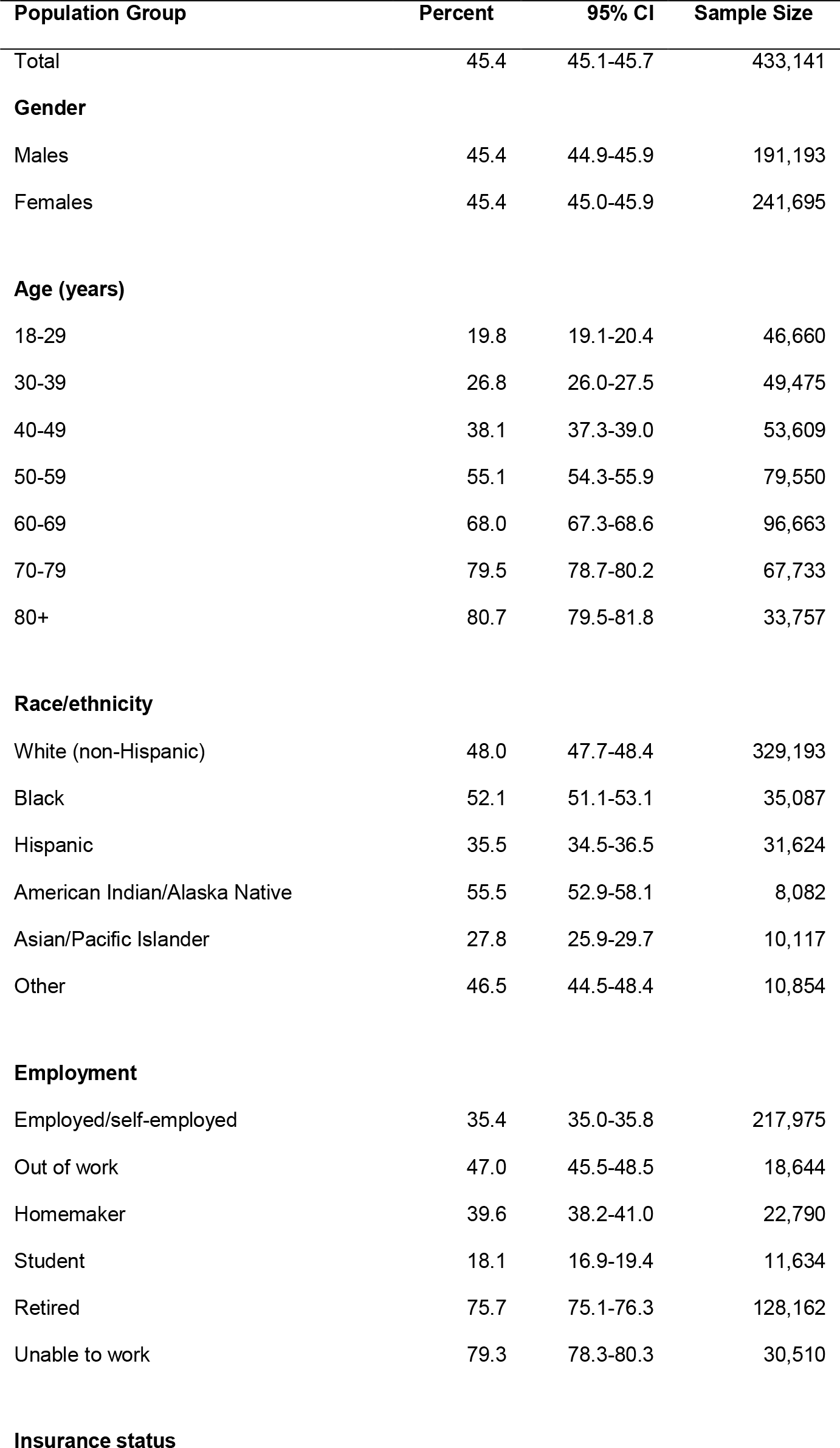
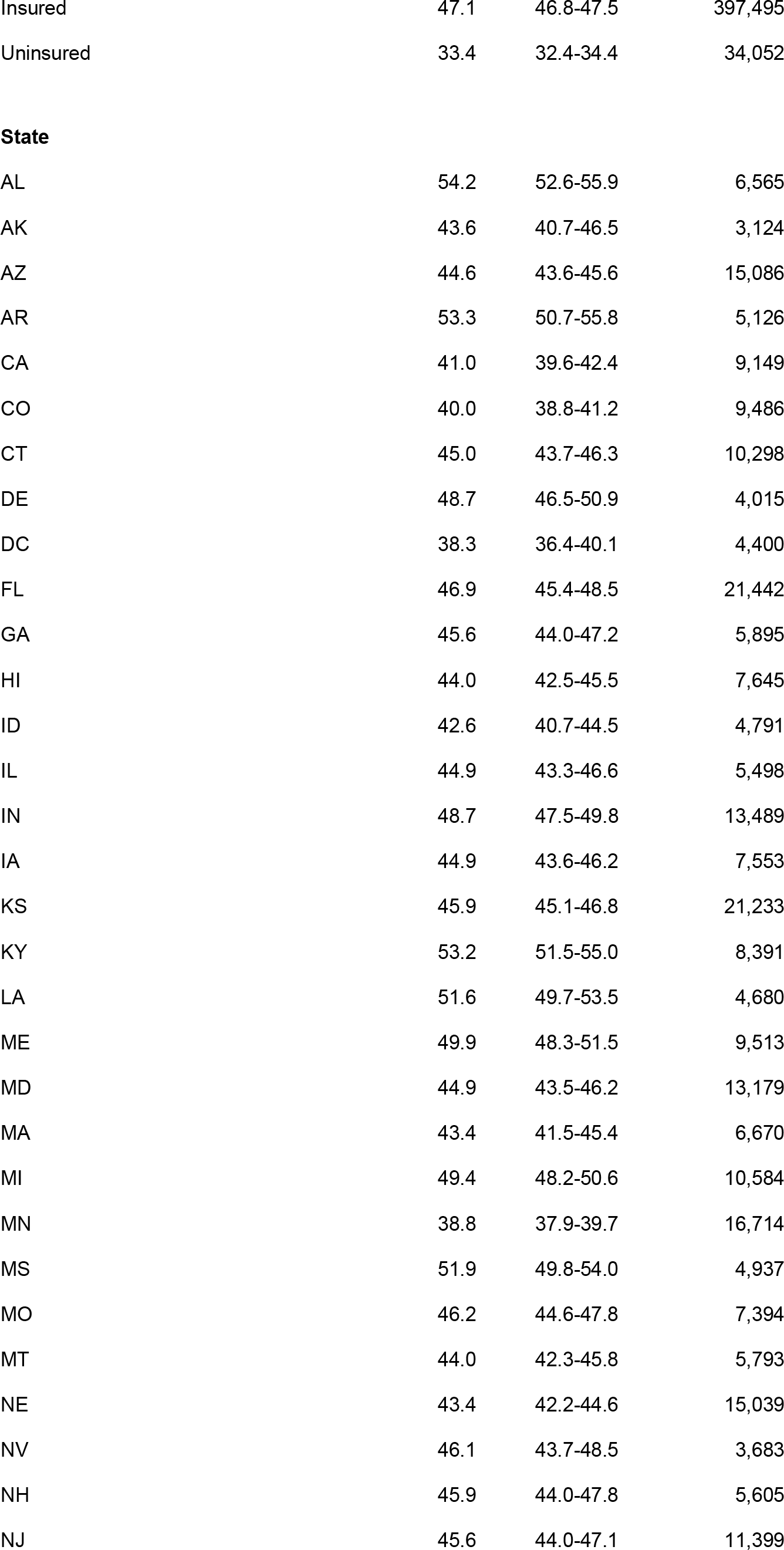
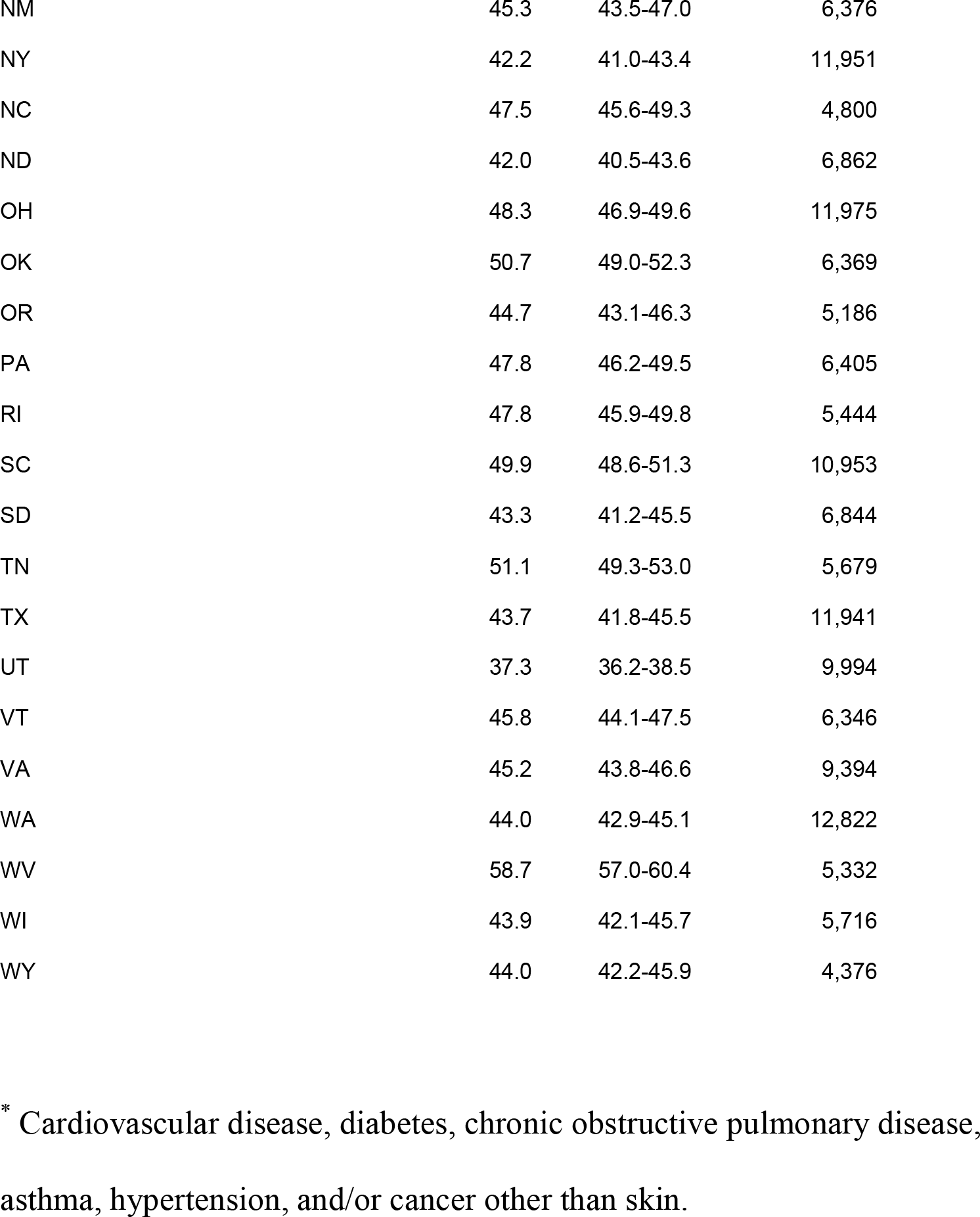
Demographics of adults with any of 6 chronic conditions ^*^ increasing risk for COVID-19 complications; 2017 Behavioral Risk Factor Surveillance System

| Population Group | Percent | 95% CI | Sample Size |
| --- | --- | --- | --- |
| Total | 45.4 | 45.1-45.7 | 433,141 |
| **Gender** |  |  |  |
| Males | 45.4 | 44.9-45.9 | 191,193 |
| Females | 45.4 | 45.0-45.9 | 241,695 |
| **Age (years)** |  |  |  |
| 18-29 | 19.8 | 19.1-20.4 | 46,660 |
| 30-39 | 26.8 | 26.0-27.5 | 49,475 |
| 40-49 | 38.1 | 37.3-39.0 | 53,609 |
| 50-59 | 55.1 | 54.3-55.9 | 79,550 |
| 60-69 | 68.0 | 67.3-68.6 | 96,663 |
| 70-79 | 79.5 | 78.7-80.2 | 67,733 |
| 80+ | 80.7 | 79.5-81.8 | 33,757 |
| **Race/ethnicity** |  |  |  |
| White (non-Hispanic) | 48.0 | 47.7-48.4 | 329,193 |
| Black | 52.1 | 51.1-53.1 | 35,087 |
| Hispanic | 35.5 | 34.5-36.5 | 31,624 |
| American Indian/Alaska Native | 55.5 | 52.9-58.1 | 8,082 |
| Asian/Pacific Islander | 27.8 | 25.9-29.7 | 10,117 |
| Other | 46.5 | 44.5-48.4 | 10,854 |
| **Employment** |  |  |  |
| Employed/self-employed | 35.4 | 35.0-35.8 | 217,975 |
| Out of work | 47.0 | 45.5-48.5 | 18,644 |
| Homemaker | 39.6 | 38.2-41.0 | 22,790 |
| Student | 18.1 | 16.9-19.4 | 11,634 |
| Retired | 75.7 | 75.1-76.3 | 128,162 |
| Unable to work | 79.3 | 78.3-80.3 | 30,510 |
| **Insurance status** |  |  |  |
| Insured | 47.1 | 46.8-47.5 | 397,495 |
| Uninsured | 33.4 | 32.4-34.4 | 34,052 |
| **State** |  |  |  |
| AL | 54.2 | 52.6-55.9 | 6,565 |
| AK | 43.6 | 40.7-46.5 | 3,124 |
| AZ | 44.6 | 43.6-45.6 | 15,086 |
| AR | 53.3 | 50.7-55.8 | 5,126 |
| CA | 41.0 | 39.6-42.4 | 9,149 |
| CO | 40.0 | 38.8-41.2 | 9,486 |
| CT | 45.0 | 43.7-46.3 | 10,298 |
| DE | 48.7 | 46.5-50.9 | 4,015 |
| DC | 38.3 | 36.4-40.1 | 4,400 |
| FL | 46.9 | 45.4-48.5 | 21,442 |
| GA | 45.6 | 44.0-47.2 | 5,895 |
| HI | 44.0 | 42.5-45.5 | 7,645 |
| ID | 42.6 | 40.7-44.5 | 4,791 |
| IL | 44.9 | 43.3-46.6 | 5,498 |
| IN | 48.7 | 47.5-49.8 | 13,489 |
| IA | 44.9 | 43.6-46.2 | 7,553 |
| KS | 45.9 | 45.1-46.8 | 21,233 |
| KY | 53.2 | 51.5-55.0 | 8,391 |
| LA | 51.6 | 49.7-53.5 | 4,680 |
| ME | 49.9 | 48.3-51.5 | 9,513 |
| MD | 44.9 | 43.5-46.2 | 13,179 |
| MA | 43.4 | 41.5-45.4 | 6,670 |
| MI | 49.4 | 48.2-50.6 | 10,584 |
| MN | 38.8 | 37.9-39.7 | 16,714 |
| MS | 51.9 | 49.8-54.0 | 4,937 |
| MO | 46.2 | 44.6-47.8 | 7,394 |
| MT | 44.0 | 42.3-45.8 | 5,793 |
| NE | 43.4 | 42.2-44.6 | 15,039 |
| NV | 46.1 | 43.7-48.5 | 3,683 |
| NH | 45.9 | 44.0-47.8 | 5,605 |
| NJ | 45.6 | 44.0-47.1 | 11,399 |
| NM | 45.3 | 43.5-47.0 | 6,376 |
| NY | 42.2 | 41.0-43.4 | 11,951 |
| NC | 47.5 | 45.6-49.3 | 4,800 |
| ND | 42.0 | 40.5-43.6 | 6,862 |
| OH | 48.3 | 46.9-49.6 | 11,975 |
| OK | 50.7 | 49.0-52.3 | 6,369 |
| OR | 44.7 | 43.1-46.3 | 5,186 |
| PA | 47.8 | 46.2-49.5 | 6,405 |
| RI | 47.8 | 45.9-49.8 | 5,444 |
| SC | 49.9 | 48.6-51.3 | 10,953 |
| SD | 43.3 | 41.2-45.5 | 6,844 |
| TN | 51.1 | 49.3-53.0 | 5,679 |
| TX | 43.7 | 41.8-45.5 | 11,941 |
| UT | 37.3 | 36.2-38.5 | 9,994 |
| VT | 45.8 | 44.1-47.5 | 6,346 |
| VA | 45.2 | 43.8-46.6 | 9,394 |
| WA | 44.0 | 42.9-45.1 | 12,822 |
| WV | 58.7 | 57.0-60.4 | 5,332 |
| WI | 43.9 | 42.1-45.7 | 5,716 |
| WY | 44.0 | 42.2-45.9 | 4,376 |

\* Cardiovascular disease, diabetes, chronic obstructive pulmonary disease, asthma, hypertension, and/or cancer other than skin.

State results presented in Table 2 list the number of adults in each state at increased risk of complications and the percentage that number represents among all US adults who report at least one of the co-morbidities. These results were obtained directly from Stata which takes into account the complex sample design so will not agree with results obtained by simply dividing the state population estimate in the Table by 111.9 million. Results for reporting a seasonal flu vaccination in the past year were 40.3% (40.0-40.6) among all adults, including 33.7% (33.3- for adults with none of the comorbidities and 48.0% (47.5-48.5) among those with any of the six.

**Table 2.** Number of adults with any of 6 chronic conditions* increasing risk for COVID-19 complications and the percentage of the total in each state; 2017 Behavioral Risk Factor Surveillance System.

| State | # Adults at risk | % of total |
| --- | --- | --- |
| AL | 1,997,864 | 1.78 |
| AK | 237,208 | 0.21 |
| AZ | 2,351,799 | 2.1 |
| AR | 1,181,105 | 1.06 |
| **CA** | 12,240,142 | 10.93 |
| CO | 1,701,776 | 1.52 |
| CT | 1,239,597 | 1.11 |
| DE | 357,530 | 0.32 |
| DC | 213,357 | 0.19 |
| **FL** | 7,696,749 | 6.88 |
| GA | 3,541,358 | 3.16 |
| HI | 486,156 | 0.43 |
| ID | 534,533 | 0.48 |
| IL | 4,404,556 | 3.93 |
| IN | 2,428,188 | 2.17 |
| IA | 1,067,133 | 0.95 |
| KS | 983,323 | 0.88 |
| KY | 1,789,444 | 1.6 |
| LA | 1,806,330 | 1.61 |
| ME | 530,809 | 0.47 |
| MD | 2,054,758 | 1.84 |
| MA | 2,302,809 | 2.06 |
| MI | 3,749,235 | 3.35 |
| MN | 1,625,778 | 1.45 |
| MS | 1,150,036 | 1.03 |
| MO | 2,137,650 | 1.91 |
| MT | 356,113 | 0.32 |
| NE | 616,905 | 0.55 |
| NV | 1,048,591 | 0.94 |
| NH | 485,340 | 0.43 |
| NJ | 3,106,880 | 2.78 |
| NM | 701,585 | 0.63 |
| **NY** | 6,419,321 | 5.73 |
| NC | 3,713,582 | 3.32 |
| ND | 243,096 | 0.22 |
| OH | 4,268,748 | 3.81 |
| OK | 1,461,941 | 1.31 |
| OR | 1,418,689 | 1.27 |
| PA | 4,738,414 | 4.23 |
| RI | 393,069 | 0.35 |
| SC | 1,912,134 | 1.71 |
| SD | 281,110 | 0.25 |
| TN | 2,610,800 | 2.33 |
| **TX** | 8,977,387 | 8.02 |
| UT | 796,721 | 0.71 |
| VT | 226,397 | 0.2 |
| VA | 2,921,171 | 2.61 |
| WA | 2,464,452 | 2.2 |
| WV | 827,193 | 0.74 |
| WI | 1,949,872 | 1.74 |
| WY | 194,544 | 0.17 |
| Total | 111,943,278 | 100 |

\* Cardiovascular disease, diabetes, chronic obstructive pulmonary disease,
asthma, hypertension, and/or cancer other than skin.

Using the data from China (3) we estimated that between 20.3% and 38.7% of Chinese adults had any of the 6 comorbid conditions with the percentage most likely between 30.5% and 34.2%.

## Discussion

We found that 45.4% of US adults, with a wide range across age groups and states, may be at heightened risk of complications from COVID-19 due to existing comorbid conditions. And while the rates increase with age group to about 80% among those ages 70 and older, due to the fact that the majority (71%) of US adults is younger than age 60, 53% of those at heightened risk of complications are also younger than age 60 years. That is consistent with the finding that >60% of adults with multiple (2 or more) chronic conditions were younger than age 65 (7). Many of these younger adults with comorbid conditions may require hospitalization for these complications. Results in Tables 1 and 2 may be helpful to localities planning healthcare system needs for COVID-19. The median value of having any of the co-morbidities for all states is 45.3, for planning purposes, states with rates in Table 1 that are above 45.3 might require proportionally more ICU beds and ventilators than those with rates below 45.3.

Currently there is a dearth of data on adults at risk for complications from COVID-19 that can be used for comparison. One source is a recent report from the Kaiser Foundation (*8*) that found 41% of US adults at risk of serious illness from COVID-19, defined as all adults ages 60 and older and those 18-59 years with heart disease, cancer, COPD, or diabetes. Note that hypertension, which is the most common comorbid condition, was omitted in that study (*8*).

From our own estimates of the fraction of adults in China with any of the 6 comorbid conditions, we estimated that fraction as likely between 30.5% and 34% of all adults based on the average case fatality rate or that of the most prevalent condition, and within the range of 20.3-38.7%. From these few comparisons, it appears that the percentage of US adults with comorbid conditions increasing their risk of complications from COVID-19 is even higher than that in China. The implications for the US, if this is true, are disturbing but would be consistent with obesity rates in China being lower than in the US (*9*). In a recent US study that included all these comorbid conditions except cancer, obesity was a major contributor to diabetes, hypertension, and asthma and at least an indirect contributor to CVD (*10*). Other risk factors also contribute to these comorbid conditions (*11*) and will have different prevalence rates in different countries and population groups within countries.

The list of chronic conditions used in our analysis is very similar to groups at increased risk for seasonal influenza complications (*12*) except that latter group includes obese adults and children <2 years and excludes people with hypertension. Both lists include people with chronic diseases for which behavioral risk factors have been well identified (*10,11*). Behavioral risk factors appear to be very common among US adults with >95% of all adults reporting one or more (*10*) of the 7 CVD risk factors of smoking, sedentary lifestyle, obesity, diabetes, hypertension, high cholesterol, and inadequate fruit and vegetable consumption. Results from that study (*10*) found those 7 risk factors together contributed to an average of 41.4% of the burden of 5 of the 6 comorbidities (all except cancer) used in this current study, with obesity and smoking contributing the most.

Results showing seasonal flu vaccination rates below 50%, even for those with any of the 6 chronic conditions that increase risk of complication from COVID-19, are concerning. Although a vaccine specific for this coronavirus is currently unavailable, results for seasonal flu vaccination suggest that it may not be widely used. It is encouraging that those with any of the 6 co-morbidities appear more likely to be vaccinated for seasonal influenza than those with none.

### Limitations

Our study does not address possible differences in contracting the disease, only the risk of developing complications among those with COVID-19, based on results from China (*1-3*). Only non-institutionalized adults are surveyed so 1.3 million adults in nursing homes (*13*) were excluded which almost certainly underestimates risk, as the first death in the US from COVID-19 was a nursing home resident (*14*). Data are self-reported and reliability and validity can vary for different measures tested (*6*). But as long as a respondent was told they had a chronic condition, validity was high. Low response rates could introduce bias but, as noted, validity appears high for the measures used in this study. Results are specific for this coronavirus in the US.

## Conclusion

These results indicating 45.4% of adults are at potentially heightened risk of complications from COVID-19 due to chronic conditions should help state health officials in estimating health care needs. The US may never determine an accurate case fatality rate but it appears likely that it would be greater than the 3.8% reported for China. It should also be clear that while adults ages 70 and older are at higher risk it is because they are more likely to have the chronic conditions identified as increasing risk but that the majority of adults at risk are actually younger than age 70.

## Data Availability

On-line with link provided in paper https://www.cdc.gov/brfss/data_documentation/index.htm

